# Dopamine transporter in obesity: a meta-analysis

**DOI:** 10.1101/2021.01.05.21249294

**Authors:** Kyoungjune Pak, Keunyoung Kim, In Joo Kim

## Abstract

The brain plays a major role in controlling the desire to eat. This meta-analysis aimed to assess the association between dopamine transporter (DAT) availability, and obesity. We performed a systematic search of MEDLINE (from inception to November 2020) and EMBASE (from inception to November 2020) for articles published in English using the keywords “dopamine transporter,” “obesity,” and “neuroimaging”. Data were plotted for each radiopharmaceutical, and linear regression was used to describe the relationship between DAT ratio, and body mass index (BMI), spline curves were adopted to fit data between DAT ratio and BMI. Five studies including 421 subjects were eligible for inclusion in this study. Two studies with ^123^I-FP-CIT, one with ^99m^Tc-TRODAT, one with ^123^I-PE21, and one with ^18^F-FP-CIT were included. DAT availabilities from ENC-DAT project were higher than those from PPMI database both for caudate nucleus, and putamen. As there might be the inter-study variability, we calculated DAT ratio, after dividing DAT availabilities of subjects with overweight/obese BMI with mean DAT availabilities of subjects with normal BMI. In conclusion, we have shown that DAT availability of subjects with overweight/obesity was not different from those with normal BMI.

## 1. INTRODUCTION

Obesity rate has nearly tripled worldwide since 1975, and has become one of the major public health threats(2), as obesity is known to be a risk factor for malignancies of colon (2), pancreas (3), thyroid (4), liver (5), and uterus (6) as well as for cardiovascular diseases (7). Obesity arises from energy intake that chronically exceeds the energy expenditure (8). This excessive eating behavior as known as overeating has phenomenological similarities with excessive drug use in addiction (9). In this regard, researchers have focused on how the brain regulates eating behavior, and how obesity affects the brain (10, 11). Among the neurotransmitters synthesized in the brain, dopamine plays a major role in eating behavior as well as the executive function, motor control, and motivation (12-14). The role of the dopamine system in obesity, and eating behavior has been investigated with the help of neuroimaging using radiopharmaceuticals, as there is no direct method to measure dopamine levels in the human brain (11). Especially, dopamine receptor (DR) has been discussed widely with regard to obesity. However, the association between obesity and DR remains unclear due to the lack of agreement between the previous studies (15-18), probably due to the characteristics of the radiopharmaceuticals, or the study design.

Dopamine transporter (DAT) is a transmembrane protein that drives reuptake of extracellular dopamine into presynaptic neurons (19). Also, DAT is a major target for various pharmacologically active drugs (19). However, DAT has not been investigated widely in the view of obesity, in addition, the association of DAT with obesity showed the inconsistent results. In the study by Chen et al., DAT availability measured from ^99m^Tc-TRODAT single-photon emission computed tomography (SPECT) was negatively correlated with body mass index (BMI) (20). However, other studies did not find the significant association between DAT availability and BMI (1, 21, 22). Therefore, we performed a meta-analysis to assess the association between DAT availability measured with radiopharmaceuticals, and obesity.

## 2. MATERIALS AND METHODS

### 2.1 Data Search and Study Selection

We performed a systematic search of MEDLINE (from inception to November 2020) and EMBASE (from inception to November 2020) for articles published in English using the keywords “dopamine transporter,” “obesity,” and “neuroimaging.” All searches were limited to human studies. The inclusion criteria were neuroimaging studies of DAT availability of striatum (striatum, caudate nucleus, putamen, ventral striatum) from healthy subjects in association with BMI. Reviews, abstracts, and editorial materials were excluded. Duplicate articles were excluded. If there was more than one study from the same institution, the study reporting information most relevant to the present study was included. Two authors performed the literature search and screened the articles independently, and discrepancies were resolved by consensus.

### 2.2 Data Extraction and Statistical Analysis

Two reviewers independently extracted the following information from the reports: first author, year of publication, country, radiopharmaceuticals, number of subjects, BMI, and DAT availability. Data were extracted by three methods; 1) BMI and the corresponding DAT availability were extracted from figures of each study using Engauge Digitizer, version 12.1 (http://digitizer.sourceforge.net), 2) the calculated DAT availability were downloaded from the shared database, 3) previously published data from our group were included. T-tests were done to compare the means of DAT availabilities from each database. Mean DAT availabilities of subjects with normal BMI (18.5≤BMI<25.0kg/m^2^) were calculated, and DAT availabilities of subjects with overweight/obese BMI (25.0kg/m^2^ or more) were divided with mean DAT availabilities of subjects with normal BMI, and defined as DAT ratio. Data were plotted for each radiopharmaceutical, and linear regression was used to describe the relationship between DAT ratio, and BMI, spline curves were adopted to fit data between DAT ratio and BMI. Data analysis was done using Prism 8 for macOS, version 8.3.1. (GraphPad Software, LLC, San Diego, CA, USA).

## 3. RESULTS

### 3.1. Study Characteristics

The electronic search identified 423 articles. Conference abstracts (n=183), animal studies (n=50), non-English studies (21), and studies that did not meet the inclusion criteria after screening the title or abstract (n=159) were excluded. After reviewing the full text of 10 articles, 5 studies including 421 subjects were eligible for inclusion in this study (1, 20, 22-24) (Figure 1). Two studies with ^123^I-FP-CIT (22, 24), one with ^99m^Tc-TRODAT (20), one with ^123^I-PE21 (1), and one with ^18^F-FP-CIT (23) were included. DAT availability was extracted from figures in 3 studies (1, 20, 22). In 3 studies that extracted data from figures, BMI and the corresponding DAT availability were extracted from 50 of 50 subjects (20), 114 of 123 subjects (22), and 33 of 33 subjects (1). The calculated DAT availability was downloaded from website of the shared database (Parkinson’s Progression Markers Initiative, PPMI; http://www.ppmi-info.org/) in one study (24). Previously published data from our group was extracted in one study (23). Three studies included the subjects from each institution (1, 20, 23), and the study by van de Giessen et al. (22) analyzed the data of European databases of [123I]FP-CIT SPECT scans of healthy controls (ENC-DAT; http://earl.eanm.org/cms/website.php?id=/en/projects/enc-dat.htm) project with 13 sites from 10 European countries without overlaps with PPMI database (24). Table 1 summarizes the study characteristics.

**Table 1.** Studies included in a meta-analysis

| Author | Year of publication | Country | Radiopharmaceutical | No. of subjects | ROI | Source of data extraction |
| --- | --- | --- | --- | --- | --- | --- |
| Chen et al | 2008 | Taiwan | $^{99m}\text{Tc}$-TRODAT | 50 | Striatum | Figure |
| van de Giessen et al | 2013 | ENC-DAT project<br>(13 sites from 10 European countries) | $^{123}\text{I}$-FP-CIT | 123 | Caudate nucleus, putamen | Figure |
| Thomsen et al | 2013 | Denmark | $^{123}\text{I}$-PE21 | 33 | Striatum | Figure |
| Pak et al | 2020 | Republic of Korea | $^{18}\text{F}$-FP-CIT | 33 | Ventral striatum, caudate nucleus, putamen | Previously published data |
| PPMI | | 33 sites | $^{123}\text{I}$-FP-CIT | 167 | Caudate nucleus, putamen | Website of the shared database |

ROI, region of interest; VST, ventral striatum; ENC-DAT, European databases of [ $^{123}\text{I}$ ]FP-CIT SPECT scans of healthy controls; PPMI, Parkinson's Progression Markers Initiative

**Figure 1.**
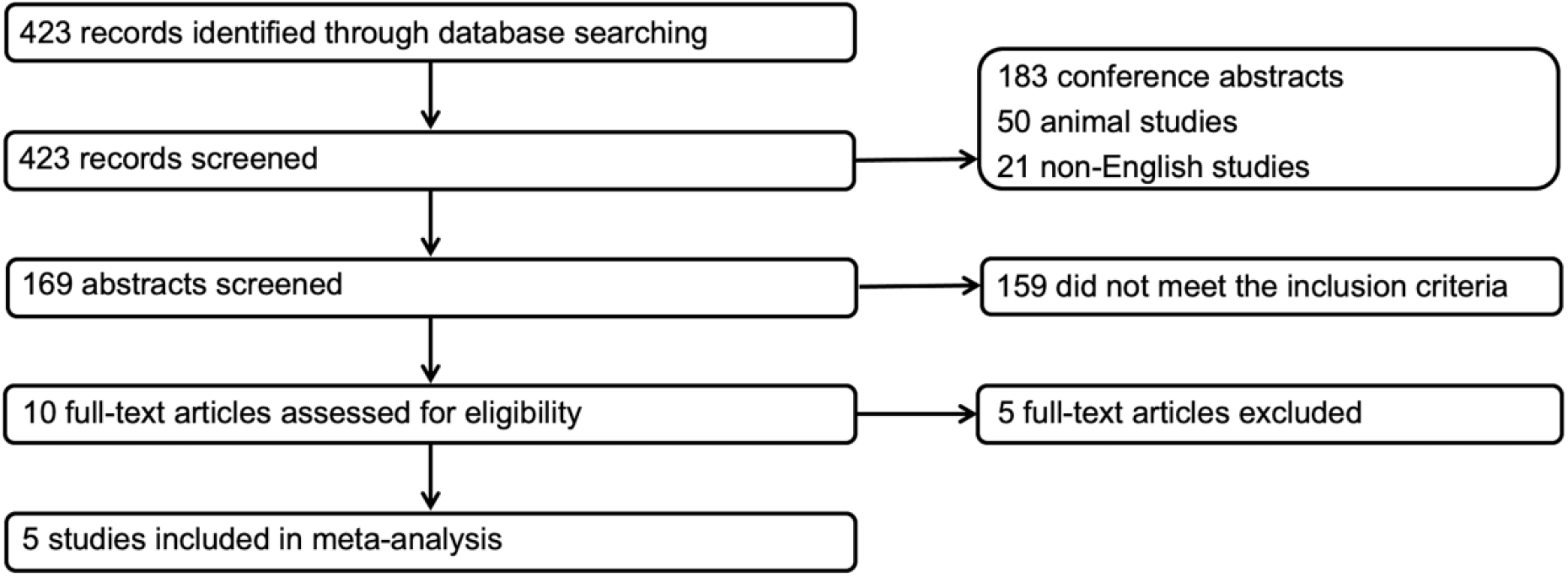
Flowchart

### 3.2. Association of DAT availability or DAT ratio with BMI

Data of 2 studies with ^123^I-FP-CIT (22, 24) were plotted for each ROI (caudate nucleus, putamen) (Figure 2). DAT availabilities from ENC-DAT project (22) were higher than those from PPMI database (24) both for caudate nucleus (p<0.0001), and putamen (p<0.0001) (Figure 2). As there might be the inter-study variability, we calculated DAT ratio, after dividing DAT availabilities of subjects with overweight/obese BMI with mean DAT availabilities of subjects with normal BMI. Linear regression between BMI, and DAT ratio was not significant for both caudate nucleus (p=0.4730), and putamen (p=0.5921). Spline curves of DAT ratio from caudate nucleus, and putamen did not increase or decrease as BMI increases (Figure 3). In the study by Chen et al. with ^99m^Tc-TRODAT, 14 subjects had BMI of 25kg/m^2^ or more (20). DAT ratio was lower than 1 in subjects with BMI of 25kg/m^2^ or more except for 1 subject, with the trend of decrease as BMI increases (Figure 4A). In the study by Thomsen et al. with ^123^I-PE21, 21 subjects had BMI of 25kg/m^2^ or more (1). Spline curves of DAT ratio from striatum did not increase or decrease as BMI increases (Figure 4B). As there were 6 subjects with BMI of 25kg/m^2^ or more in the study by Pak et al., curve fitting for DAT ratio, and BMI could not be done (23).

**Figure 2.**
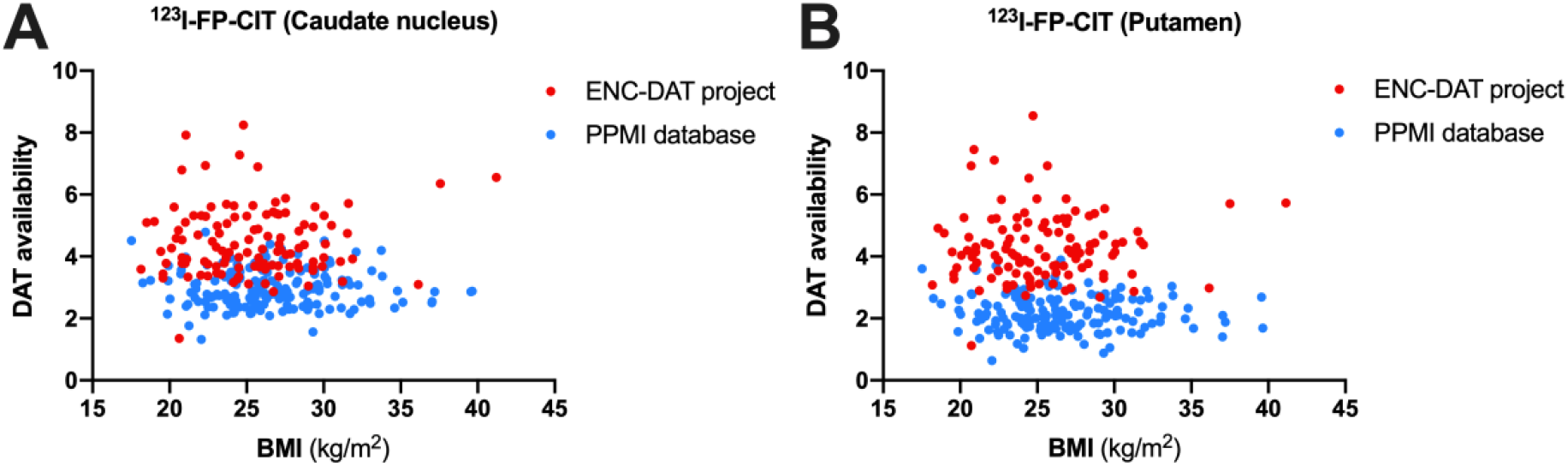
Scatter plots of DAT availability and BMI; A. Caudate nucleus; B. Putamen

**Figure 3.**
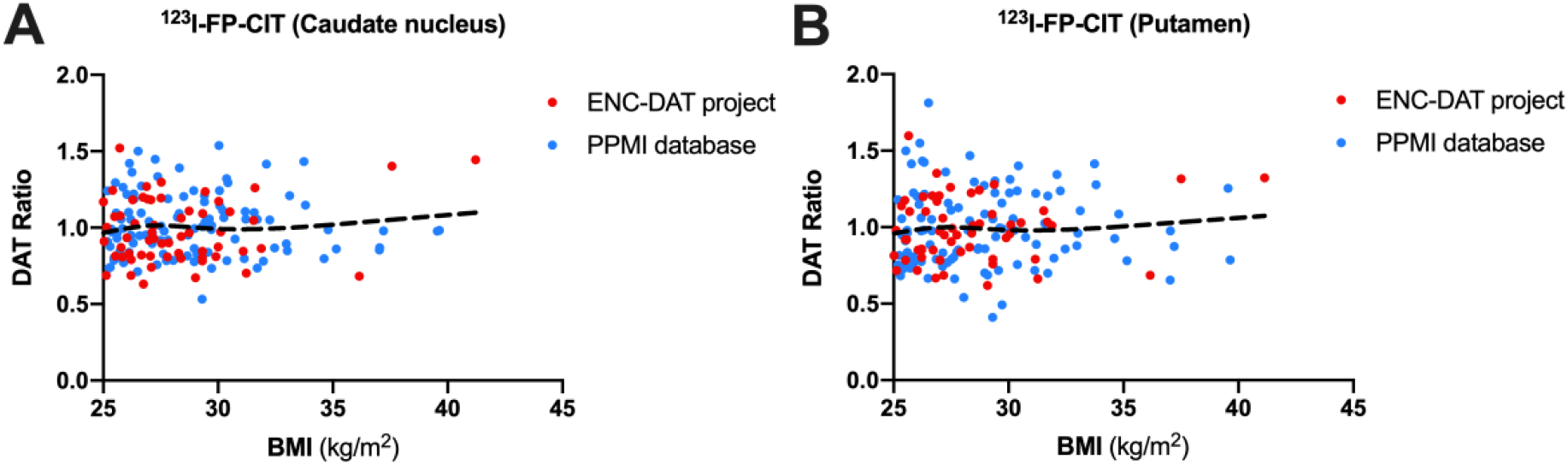
Scatter plots and spline curves of DAT ratio and BMI; A. Caudate nucleus; B. Putamen

**Figure 4.**
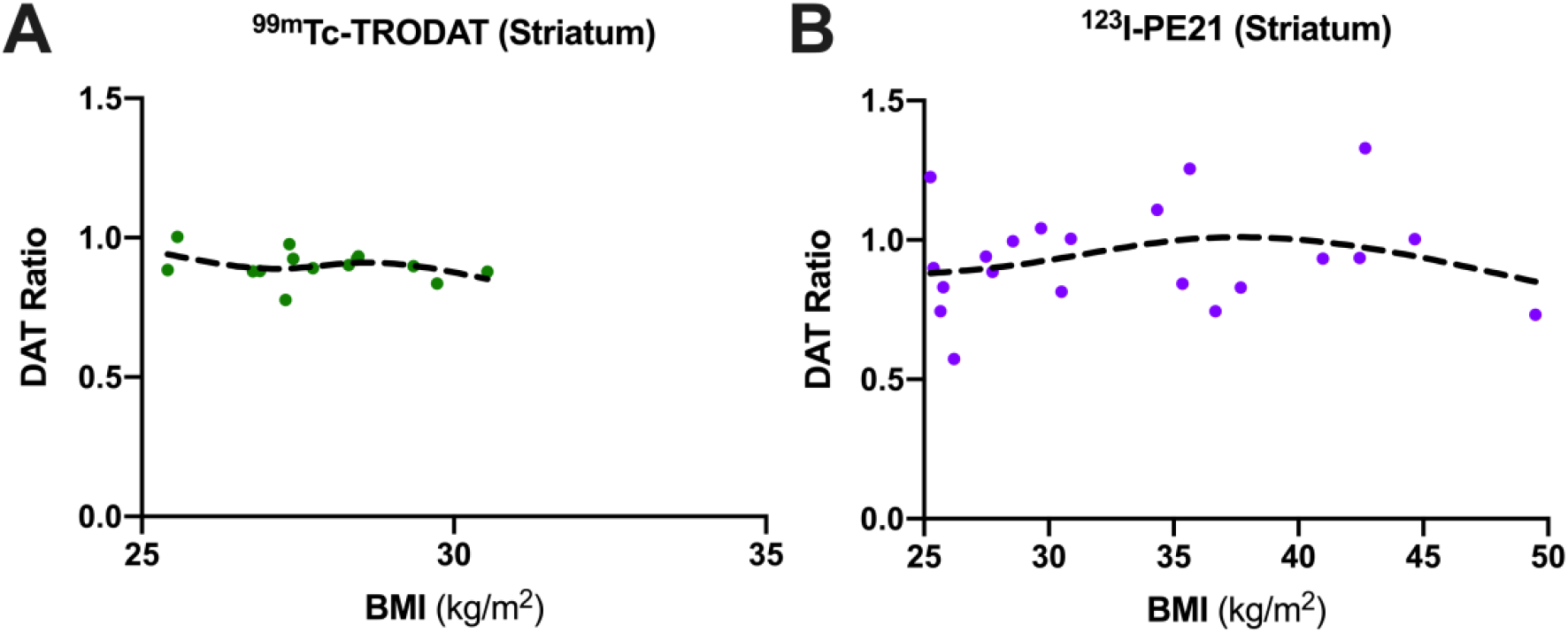
Scatter plots of DAT ratio, and BMI; A. ^99m^Tc-TRODAT; B. ^123^I-PE21

## 4. DISCUSSION

In this study, we have shown that DAT availability of subjects with overweight/obesity was not different from those with normal BMI. Although DAT has not been discussed widely with regard to obesity, 5 studies of 421 subjects could be included in this study. Except for one study by Chen et al.(20), 4 studies have shown no significant association between DAT availability and BMI (1, 21-23). Chen et al. used ^99m^Tc-TRODAT SPECT to measure DAT availability, and showed the negative correlation between DAT availability and BMI (20). ^99m^Tc-TRODAT is a radiopharmaceutical based on a tropane structure, showing similar diagnostic performance with ^123^I-FP-CIT to detect Parkinsonism (25). However, there might be the difference of binding capacity of radiopharmaceuticals in healthy subjects. Among 3 other studies that included in this meta-analysis, 2 studies with ^123^I-FP-CIT (22, 24), and 1 study with ^123^I-PE21 (1) did not show the significant correlation between DAT availability and BMI. In a previous study by Ziebell et al.(26), ^123^I-FP-CIT and ^123^I-PE21 showed the same target-to-background ratios, however, ^123^I-FP-CIT gave rise to 3-fold higher counting rates per injected dose. From 2 studies with ^123^I-FP-CIT, we extracted the data from the figure using the software (22), or the website of PPMI (24). In both of 2 studies, DAT availability was measured from striatal regions of caudate nucleus, and putamen (22, 24). However, as shown in figure 2, systematic difference between 2 studies might exist due to study design such as image acquisition, study population. Therefore, we plotted the data of DAT ratio of overweight/obese subjects to investigate the DAT availability adjusted for inter-study variability.. In the study by Thomsen et al.(1), ^123^I-PE21 also did not change significantly according to BMI. However, the numbers of overweight/obese subjects included in studies of Chen et al. with ^99m^Tc-TRODAT (20), and Thomsen et al.(1) with ^123^I-PE21 were small.

Dopamine plays a major role in modulating motivation and in reward processing (27). There are two major hypotheses regarding the role of dopamine. The first hypothesis, dopamine hyperresponsiveness, explains that there is a hypersensitivity to rewards which is related to an increased behavioural salience towards food leading to the excessive intake of highly palatable foods(28, 29). The second hypothesis, reward deficiency syndrome, states that subjects who are insensitive to rewards overeat to increase their endogenous dopamine levels (28, 29). Therefore, a nonlinear relationship of an inverted parabola has been proposed between the DR and BMI based on previous study (30). In mild obesity, the change in DR is not significant. However, with the onset of moderate obesity, the responsiveness of dopamine increases until the BMI rises to approximately 35 to 40 kg/m^2^, following which a reward deficiency occurs, in severe obesity (28, 30). Brain dopamine neurotransmission is regulated by DAT, which drives reuptake of extracellular dopamine into presynaptic neurons (19). Previously, we used the glucose infusion to reflect food intake, and we have discovered that intravenous glucose loading induced substantial increases of striatal DAT availability by the action of insulin (23). From animal studies, insulin acts on striatal insulin receptors to enhance the surface expression of DAT through the PI3 kinase signalling pathway (31, 32). Therefore, food intake induced insulin signal works on the reward system through the dopamine function (32, 33).

In this study, we investigated the association of DAT availability with overweight/obesity, not with eating behavior. The state of overweight/obesity might be the results of excessive repetition of eating behavior. Therefore, the action of DAT on eating behavior might be different from that of the state of overweight/obesity. DAT is a major target for various pharmacologically active drugs. However, DAT is insensitive to the concentration of synaptic dopamine, as no change of DAT availability was observed after depletion of dopamine or release of dopamine, different from DR (34, 35). Therefore, no significant change of DAT in overweight/obesity may result from insensitivity of DAT to synaptic dopamine concentration, although synaptic dopamine concentration is increased or decreased in overweight/obesity. In addition, DAT may not have a role in the state of overweight/obesity, although it has an important role through the action of insulin in eating behavior.

This study has several limitations. First, a small number of studies were included in this meta-analysis. Therefore, the data from two studies of ^123^I-FP-CIT could be pooled to analyze the role of DAT in obesity. Second, radiopharmaceuticals, image processing, and data analysis techniques are different across the studies included in this meta-analysis, which might impact DAT availability. However, to adjust the inter-study variability, we adopted DAT ratio, rather than DAT availability. Third, although two authors extracted the BMI and corresponding DAT availability from figures in the studies, the data may be inaccurate.

In conclusion, dopamine plays a main role in the reward system with regard to obesity. Compared to subjects with normal BMI, those of overweight/obesity did not have a different DAT availability.

## Data Availability

DATA area available from the corresponding author on reasonable request

## ACKNOWLEDGEMENT

No

## AUTHOR CONTRIBUTION

Kyoungjune Pak; study design, write the manuscript, image analysis, statistical analysis Keunyoung Kim; image analysis

In Joo Kim; image analysis, statistical analysis

## DATA AVAILABILITY

The datasets generated during the current study are available from the corresponding author on reasonable request.

